# Access to internet, smartphone usage, and acceptability of mobile health technology among cancer patients

**DOI:** 10.1101/19012823

**Authors:** Rashmika Potdar, Arun Thomas, Matthew DiMeglio, Kamran Mohiuddin, Djeneba Audrey Djibo, Krzysztof Laudanski, Claudia M. Dourado, John Charles Leighton, Jean G. Ford

## Abstract

**Purpose:** The use of mobile health (mHealth) technologies to augment patient care, enables providers to communicate remotely with patients enhancing the quality of care and patient engagement. Few studies addressed barriers to its implementation, especially in medically underserved populations.

**Methods:** A cross-sectional study of 151 cancer patients was conducted at an academic medical center in the United States. A trained interviewer performed structured interviews regarding the barriers and facilitators of patients’ current and desired utilization of technology for healthcare services.

**Results:** Of the 151 participants, 35.8% were male and ages ranged from 21-104 years. Only 73.5% of participants currently have daily access to internet, and 68.2% currently own a smartphone capable of displaying mobile applications. Among all participants, utilization of a daily mHealth application was significantly higher in patients with a college-level degree (OR; 2.78, *p*<0.01) and lower in older patients (OR; 0.05, *p*<0.01). Differences in utilization when adjusted for current smartphone use and daily access to internet were nonsignificant. Among smartphone users, the desire to increase cancer knowledge was associated with a higher likelihood of utilizing a mHealth application (OR; 261.53, *p<*0.01).

**Conclusion:** The study suggests the access to mobile technology is the predominant determinant of utilization. Healthcare organizations should consider these factors when launching patient engagement platforms.

## 1. Introduction

As mortality rates continue to decrease among cancer patients in the United States, health care systems are faced with a growing population with complex medical, psychological, and social needs **(1)**. This has caused comprehensive care that extends years beyond treatment initiation. However, lapses in care continue to persist and much progress is needed to improve quality among cancer survivors **(2)**. Mobile health (mHealth) technology has served as a tool that can potentially address these needs and encourage active participation among patients in a variety of care settings including cancer-related care **(3–5)**.

The integration of mobile health (mHealth) applications in cancer care have been widely discussed and proposed roles include augmenting clinical processes, managing chemotherapy-related side effects, and supporting drug adherence **(6**,**7)**. The current role of mHealth in oncology has been limited to self-management, and the quality of cancer-related mobile applications varies drastically **(8**,**9)**. A recent study found that while the acceptance of mHealth technology was high among cancer patients, adoption of such technology is low. Factors predictive of the use of mHealth in this population included rural residency and a new diagnosis of cancer, indicating that geography and education are driving motivators for mHealth utilization **(10)**.

Mobile health has also been proposed as a potential tool to reduce health disparities among underserved populations through improved health literacy, medication adherence, and clinical outcomes **(11)**. Low health literacy has been associated with several poor health outcomes and partially explains the racial and socioeconomic disparities in care **(12**,**13)**. While the data regarding the efficacy of implementing mHealth initiatives are low quality and scarce, the current data that exists is promising **(5**,**8)**. A recent meta-analysis of mHealth-based interventions among breast cancer patients found that interventions consistently improved quality-of-life measures among these patients, albeit the data on the psychological effect of these interventions is less conclusive **(14)**.

However, another meta-analysis reported in pain, fatigue, and other psychological outcomes among cancer survivors utilizing mHealth-based interventions **(15)**. This suggests that the data on the efficacy of mHealth on both cancer survivorship and those undergoing treatment is still an emerging and exciting area of research.

This purpose of this study was to investigate factors associated with a willingness to utilize a mHealth application for general health maintenance among a cohort of cancer patients at an academic medical center that services a very socioeconomically diverse and medically under-served population. This study was initially launched as a quality improvement project seeking new patient engagement platforms. We hypothesized that while the acceptability of a mHealth application for cancer-related health information will be quite high, it will be limited by education-related factors and current technology use patterns. Specifically, we believe that (1) patients without current daily internet will be less accepting of a mHealth application and (2) patients that currently lack a smartphone will be less accepting of a mHealth application than their counterparts with this technology.

## 2. Materials and Methods

### 2.1 Study Design and Participants

This is a cross-sectional study of cancer patients attending the outpatient clinic and infusion center at an academic medical center in Philadelphia. The target population was adults with Inclusion criteria consisted of (1) age greater than 18 years, (2) diagnosis of malignancy, and (3) willingness to participate. Before administration, the survey was evaluated by the institutional review board (IRB) at the host institution and was deemed IRB exempt. Data was collected over a 1-month period in 2018.

### 2.2. Survey Development

This survey was designed by one of the authors (R.P.). The survey consisted of basic demographic information (age, gender, highest education, and marital status) followed by a series of questions regarding access to the internet and mobile technology. Previous use of mHealth applications as well as desired use of mHealth applications was also evaluated. All questions were arranged in yes/no format for simplicity. Also, the participants were asked to answer the questions irrespective of current technology use (*i*.*e*. using a mobile application on a smartphone in participants currently without a smartphone).

The survey was initially tested among a pilot population of 76 participants and subsequently expanded to a target of 150 participants. The sample size was determined using an a priori power analysis estimating a 20% mean difference in mHealth acceptability based on educational achievement with a Type I error rate of 0.05% and Type II error rate of 20%. Analysis indicated that 99 patients would be required to detect this difference.

### 2.3 Data Collection

Eligible participants were approached and given explanations regarding the purpose, significance, and methods of the study. After receiving consent, a trained interviewer performed structured interviews using the survey as a guide. Study data were collected and managed using Research Electronic Data Capture (REDCap) tools hosted at the study site **(16)**.

### 2.4 Statistical Analysis

The data were analyzed using Statistica® Version 13.0 (TIBCO Software Inc.) and Graphpad® PRISM 6.0 (Graphpad Software Inc.). Demographic variables were characterized by descriptive statistics. Chi-Square test for association was utilized for categorical variables. Statistical significance was defined as p < 0.05 on a two-tailed distribution. Finally, adjusted odds ratios from logistic regression analysis were presented to identify the effects of demographic information and survey answers on a patient’s willingness to utilize an application for monitoring their health condition. Adjusted odds ratio reported as (OR; mean, CI_95%_, *p-*value).

## 3. Results

A total of 151 participants were approached and all participants consented to the survey (100% response rate). The demographic information of participants is summarized in Table 1. The mean age of participants was 62.3±14.4 years of age and consisted of 35.8% male. Race demographics were not collected after the pilot population was expanded, but the pilot population consisted primarily of minority groups (83.1%). While 60.3% of participants had a high school degree or less, 39.7% obtained at least an undergraduate degree with 5.9% of total participants achieved additional education (Table 1).

**Table 1.**
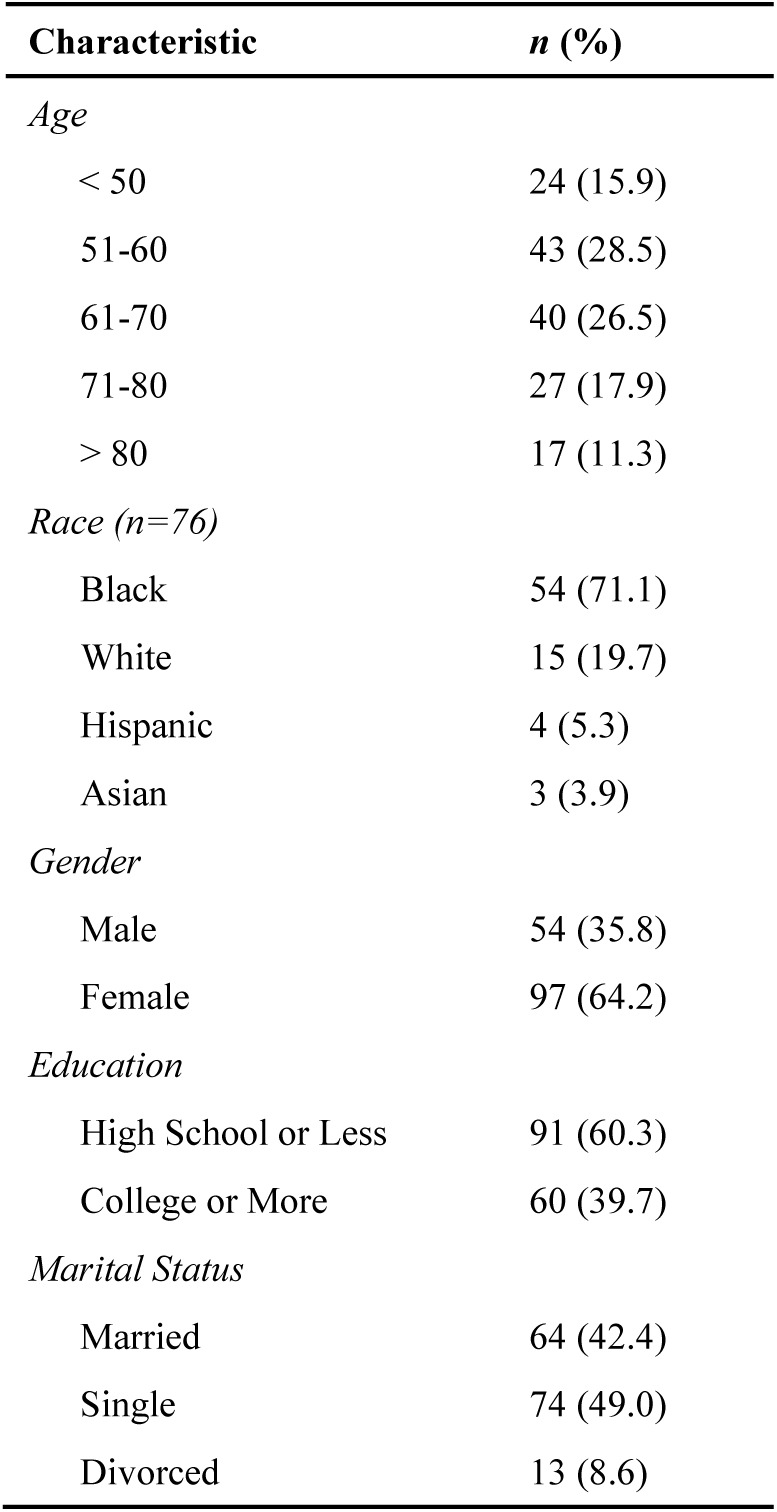
Demographic Information of Participants *(N = 151)*

The response distribution of selected survey items is outlined in Table 2. Interestingly, only 73.5% of survey respondents have daily access to internet services. Individuals without internet tended to be older (mean age of 71.9±11.2 *vs*. 58.8±13.9) and less educated (12.5% with a college degree *vs*. 49.5%) than their counterparts with internet. Similar characteristics were seen in individuals that lacked either a mobile phone or smartphone (Table 2). Among the 106 individuals that currently own a smartphone, 60.4% use phone that operates iOS software (iPhone®) and 39.6% use a phone that operates Android software (Samsung®, LG®, etc.). The frequency of previous, defined as over the past 6 months, mobile access to general health and personal health information was similar (53% and 51.7% respectively). However, 59 participants (39.1% of total) neither accessed general nor personal health information while 66 participants (43.7% of total) utilized both forms of health technology (Table 2).

**Table 2.** Response Distribution of Selected Survey Items

| Item # | Survey Item |  | n | (%) |
| --- | --- | --- | --- | --- |
| 1 | *Do you currently have daily access to the Internet?* | Yes | 111 | (73.5) |
|  |  | No | 40 | (26.5) |
| 2 | *Do you currently own a mobile phone?* | Yes | 127 | (84.1) |
|  |  | No | 24 | (15.9) |
| 7 | *Do you currently own a smartphone?* | Yes | 103 | (68.2) |
|  |  | No | 48 | (31.8) |
| 10 | *In the last six months, have you used your phone to access general health information?* | Yes | 80 | (53.0) |
|  |  | No | 71 | (47.0) |
| 11 | *In the last six months, have you used your phone to access your health care information (e.g. to schedule appointments)?* | Yes | 78 | (51.7) |
|  |  | No | 73 | (48.3) |
| 12 | *Would you want to be able to access general information related to your health/cancer via your smartphone?* | Yes | 88 | (58.3) |
|  |  | No | 63 | (41.7) |
| 13 | *Would you want to receive text messages on your phone related to your health/cancer from your doctor's office?* | Yes | 93 | (61.6) |
|  |  | No | 58 | (38.4) |
| 14 | *Would you want to use your phone to help track your cancer related information via an application or "app" on your smartphone?* | Yes | 79 | (52.3) |
|  |  | No | 72 | (47.7) |
| 15 | *Would you download an application or "app" to your phone to help increase your cancer-related knowledge?* | Yes | 87 | (57.6) |
|  |  | No | 64 | (42.4) |
| 16 | *Would you be willing to use an application or "app" on your phone daily to help monitor your health condition?* | Yes | 81 | (53.6) |
|  |  | No | 70 | (46.4) |

Future-oriented survey items (items 12 through 16) evaluated different aspects of integrating mHealth technology into practice and were met with modest support overall. However, acceptance was significantly different based on current technology utilization. Among individuals without access to the internet, the mean acceptance rate of future-oriented items was 7.5%. This pattern was also demonstrated in individuals lacking a mobile phone (4.2%) and smartphone (2.3%) (data not shown).

Regarding demographic factors influencing the willingness to utilize a mobile application for daily health monitoring, acceptance was significantly associated with both age and educational achievement (table 3). Willingness to utilize a mHealth application was highest in individuals <50 years of age (83.3% favorable) and lowest in individuals >80 years of age (82.4% unfavorable). Logistic regression revealed that age groups 61-70 (OR; 0.24, 0.07-0.90, *p*<0.01), 71-80 (OR; 0.05, 0.01-0.23, *p*<0.01), and >80 years (OR; 0.04, 0.01-0.22, *p*<0.01) were significantly less likely to utilize a daily mHealth application than individuals <50 years (Figure 1A). 71.7% of individuals with at least a college degree were favorable of a mHealth application compared to 41.8% of individuals without a college education (OR; 2.78,1.25-5.88, *p*<0.01) (Table 3 & Figure 1A). No significant differences in race, gender, or marital status were identified. However, differences among age groups and education were eliminated when adjusting for current smartphone use (Figure 1B).

**Table 3.** Comparison of demographic information to the willingness to use an application for daily health monitoring (Survey Item #16)

| Characteristic | Willing to Utilize Application<br>*n* (%) | Not Willing to Utilize Application<br>*n* (%) | Chi-square test of independence<br>($\chi^2$) |
| --- | --- | --- | --- |
| *Age* |  |  |  |
| < 50 | 20 (83.3) | 4 (16.7) | $\chi^2$ (4) = 39.7<br>*p* < 0.01 |
| 51-60 | 29 (67.4) | 14 (32.6) |  |
| 61-70 | 23 (57.5) | 17 (42.5) |  |
| 71-80 | 6 (22.2) | 21 (77.8) |  |
| > 80 | 3 (17.7) | 14 (82.4) |  |
| *Race (n=76)* |  |  |  |
| Black | 24 (44.4) | 30 (55.6) | $\chi^2$ (3) = 2.86<br>*p* = 0.41 |
| White | 5 (33.3) | 10 (66.7) |  |
| Hispanic | 3 (75.0) | 1 (25.0) |  |
| Asian | 2 (66.7) | 1 (33.3) |  |
| *Gender* |  |  |  |
| Male | 30 (55.6) | 24 (44.4) | $\chi^2$ (1) = 0.12<br>*p* = 0.73 |
| Female | 51 (52.6) | 46 (47.4) |  |
| *Education* |  |  |  |
| High School or Less | 38 (41.8) | 53 (58.2) | $\chi^2$ (1) = 13.0<br>*p* < 0.01 |
| College or More | 43 (71.7) | 17 (28.3) |  |
| *Marital Status* |  |  |  |
| Married | 35 (54.7) | 29 (45.3) | $\chi^2$ (2) = 0.05<br>*p* = 0.97 |
| Single | 39 (52.7) | 35 (47.3) |  |
| Divorced | 7 (53.9) | 6 (46.1) |  |

**Figure 1.**
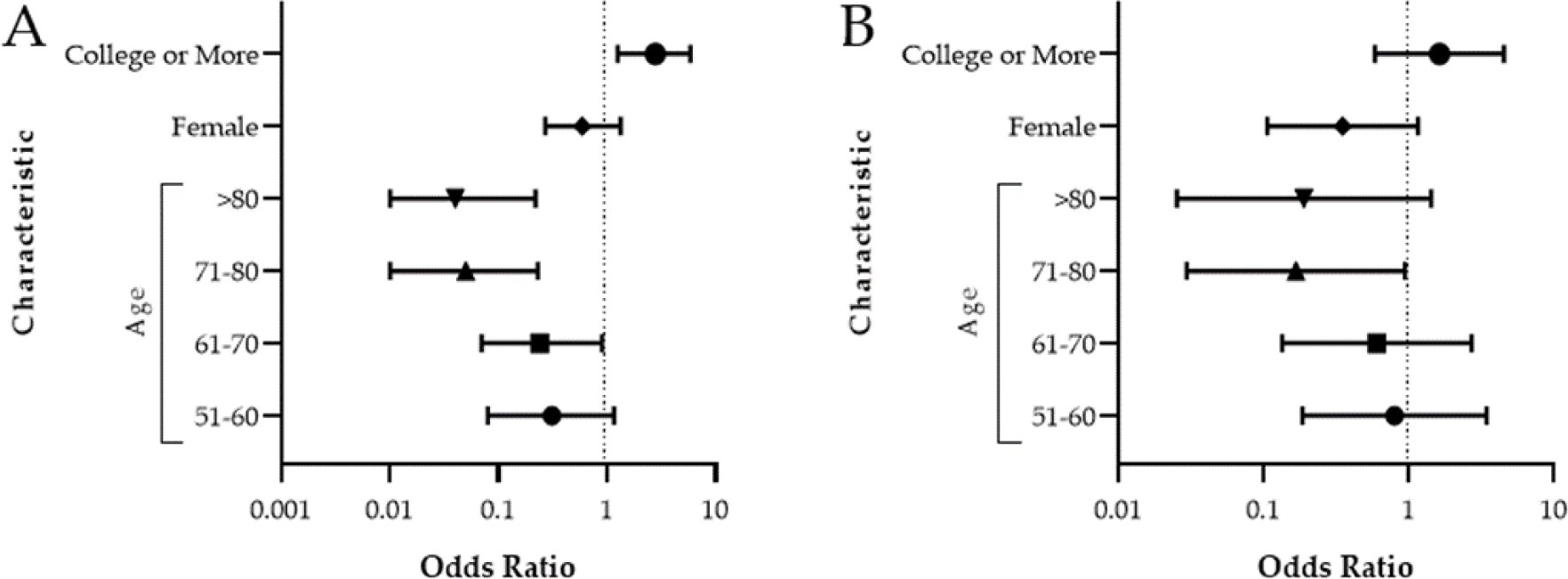
Factors associated with willingness to utilize a daily mobile application for health monitoring among all participants (A) and current smartphone users (B) reported as odds ratio. Reference group for education, gender, and age was high school or less, male, and <50 years respectively.

A desire to increase cancer-related knowledge was associated with an increased odds of utilizing a daily mHealth application (OR; 261.5, 10.13 – 6748.71, *p*<0.01). Interestingly, no other features of a mobile application exhibited signficant associations despite being similar in nature (Figure 2B). Evaluation of the frequency of each response revealed that of the 81 participants that were willing to use a daily mobile application, 80 (98.8%) also desired an application to increase cancer-related knowledge. Conversely, of the 70 individuals not willing to utilize a daily application, 63 (90.0%) also did not desire an application to increase their cancer-related knowledge (data not shown).

**Figure 2.**
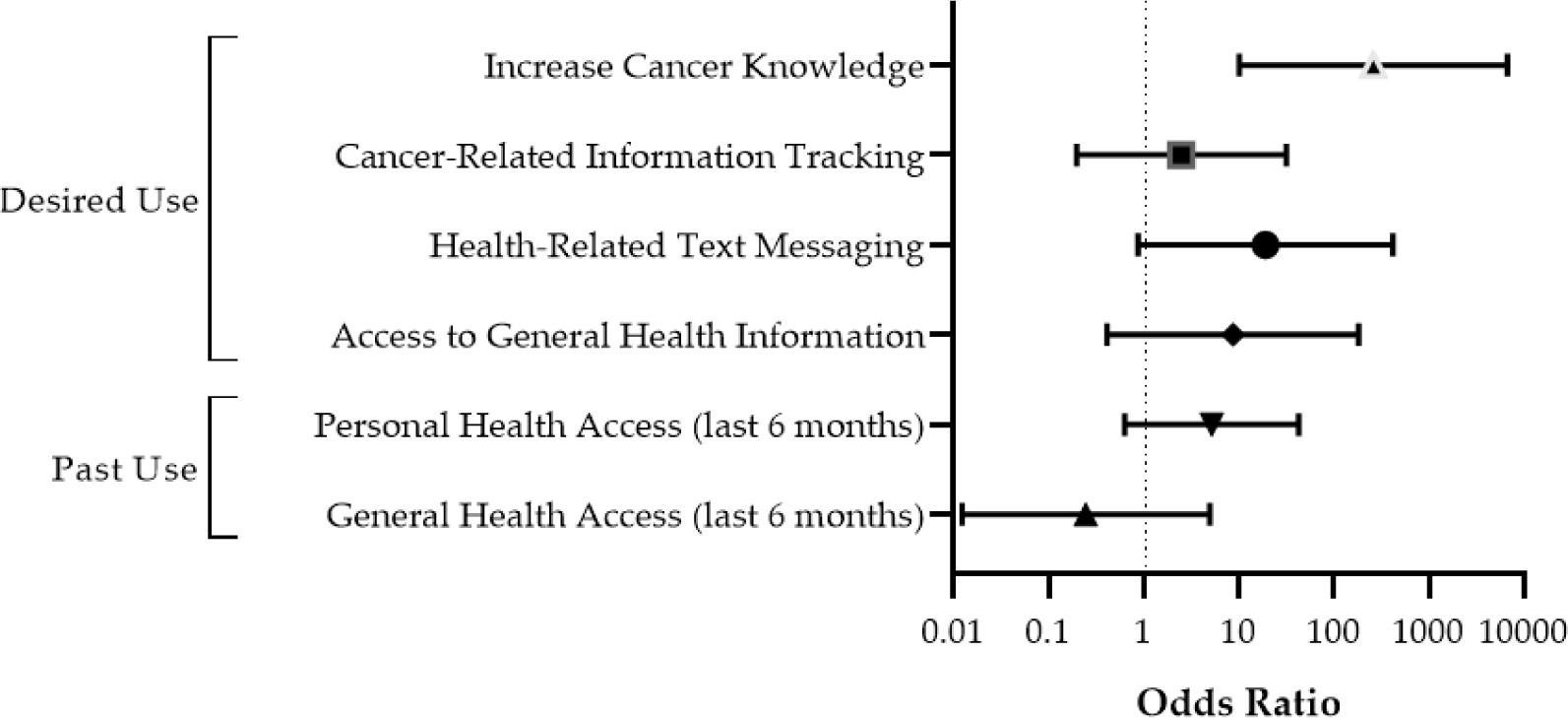
Willingness to utilize a daily mHealth application among smartphone users based on previous and desired mHealth formats reported as odds ratios. Reference value for each activity was the negative response (no).

## 4. Discussion

This study aimed to investigate the factors associated with the willingness to utilize a mHealth application among a population of cancer patients in a socioeconomically diverse and medically underserved community. Our study revealed that while the acceptability of mHealth technology was hindered by age and education-related barriers among all participants, these factors were eliminated when evaluating current smartphone users and those with daily access to internet. This suggests that past technology use is highly indicative of adoption of future technology such as a daily mobile application for cancer-related health. These findings also indicate that access to technology facilitates health-promotion activities among a predominate minority and socioeconomically diverse population.

Several studies have identified gaps in internet access along sociodemographic boundaries such that individuals with lower income, lower education, and identify as a racial minority are less likely to have access to internet services **(17**,**18)**. These findings, coupled with the abundant literature linking the prevalence of inadequate health literacy and poor clinical outcomes among African-Americans, further highlight the need for mHealth applications among minority populations in the United States **(11**,**12**,**19)**.

Our study both confirms and expands upon the previous literature. Previous literature has reported both high acceptability and positive outcome measures among cancer patients utilizing mHealth-based interventions **(9**,**10**,**14**,**20)**. However, our study has investigated both the acceptability and feasibility of mHealth-based interventions among a population of predominately racial minorities and of lower socioeconomic status. While our study reported an overall acceptance of a mHealth application for health information among cancer patients, we found that our population faces technology-related barriers at a higher prevalence than the rest of the United States. The rate of daily internet access among our patient (73.5%) is much lower than a recent Pew Research estimate of 90% **(21)**. Our population also had a much lower rate of both a mobile phone (84.1%) and smartphone ownership (68.2%) than a recent national estimate of 96% and 81% respectively **(22)**. This highlights a concern regarding implementation of smartphone-based patient engagement platforms because a large percentage of patients would be excluded from participation. Considering the existence of many cheap smartphones within the United States, future studies should investigate both monetary and non-monetary factors related to smartphone adoption among low-income communities.

We found that the desire to increase cancer knowledge was significantly associated with an increased likelihood of utilizing a daily mHealth application for cancer-related information among current smartphone users. Interestingly, we found that past mHealth-related use patterns (accessing personal or general health information on a smartphone) was not associated with the willingness to utilize a daily mHealth application. This suggests that although current smartphone users may not have accessed personal or general health information in the past, future mHealth applications that integrate cancer education with patient health information shows promise in terms of patient acceptance. This offers an exciting avenue for future patient engagement initiatives.

There are several limitations in this study. This study investigated only a small subset of patients within a population of predominately low socioeconomic status, so the results are likely not generalizable to the overall population. While a *post hoc* power analysis indicated that our study was adequately powered to detect the observed differences, there is always the possibility of detecting a difference that is not present in the overall population given our sample size. Our study also only evaluated the willingness of a theoretical mobile application, therefore further research is required to develop and test a mobile application within this population.

## 5. Conclusions

The potential use of mHealth-based interventions in a socioeconomically diverse population offers several barriers and opportunities. The relatively lower rates of daily internet access and smartphone use in a low-income patient population appear to be the largest barrier to mHealth acceptance, while education and age-related barriers are eliminated when adjusting for current smartphone use. The desire to increase cancer knowledge was significantly associated with an increased likelihood to utilize a daily mHealth application and offers an opportunity to integrate patient education with future mHealth-based interventions.

## Data Availability

Data will be made accessible upon reasonable request and in accordance with institutional review board data transfer guidelines.

## Supplementary Materials

Supplementary File 1. Patient Questionnaire

## Author Contributions

Conceptualization, study design, and methodology, R.P., K.M., C.D., J.L., and J.F; data analysis, M.D. and K.L.; writing – original draft preparation, M.D. and K.L.; writing – reviewing and editing, all authors participated.

## IRB Statement

The study was approved by the institutional ethics committee at the host institution and was conducted in concordance with the tenets of the Declaration of Helsinki

